# Recent and convergent reversion to serotype Ogawa in the AFR12 sublineage of *Vibrio cholerae* O1 El Tor in Cameroon

**DOI:** 10.1101/2025.04.25.25326187

**Authors:** Pierrette Landrie Simo Tchuinte, Rosanne Minome Ngome, Elisabeth Njamkepo, Larissa Diane Tagne Magne, William Mbanzouen, Marcelle Abanda, Esther Sokeng, Laurence Noubissi-Jouegouo, Flaubert Tassadjo, Manuella Ango, Cédric Thierry Roland Fouda, Urbaine Ngon, Ariane Nzouankeu, Oumar Aïcha, Ronald Perraut, Yves Eric Donan Denon, Honoré Bankolé Sourou, Halatoko Afiwa Wemboo, Adodo Yao Sadji, Fati Sidikou, Caroline Rouard, Marie-Laure Quilici, Jean-Marc Collard, François-Xavier Weill

## Abstract

Since 1971, Cameroon has experienced several outbreaks of cholera due to strains of the seventh pandemic *V. cholerae* O1 El Tor (7PET) lineage originating from South Asia. Over the last six years, more than 26,000 cholera cases have been reported. The aim of this study was to perform a genomic characterisation of the *V. cholerae* O1 isolates collected during recent cholera outbreaks in Cameroon. We investigated the virulence, antimicrobial resistance (AMR) signatures and phylogenetic relationships between 164 clinical *V. cholerae* isolates representative of the successive outbreaks that were collected between 2018 and 2023. A phylogenomic analysis of more than 1,700 7PET genomes —including 45 from isolates collected in Cameroon between 1971 and 2011 — was performed to place the recent 7PET strains in a broader phylogenetic context. We found that all the recent Cameroonian isolates studied belonged to genomic wave 3 of the 7PET lineage. They clustered together within the AFR12 sublineage (which was identified for the first time in Cameroon in 2009 and with contemporary isolates from other countries in the same region). Serotyping and genome analysis revealed a reversion from serotype Inaba (all isolates between 2018 and 2019) to Ogawa (all isolates between 2021 and 2023) in two different strains originating from different regions of Cameroon. Finally, the AFR12 isolates studied here were less resistant to antimicrobial drugs than the AFR12 isolates identified in Cameroon between 2009 and 2011 due to a 10-kb deletion in the integrative conjugative element conferring multidrug resistance, ICE*Vch*Ind5, resulting in the loss of four antimicrobial resistance (AMR) genes (*strA*, *strB*, *floR* and *sul2*). Our findings confirm the complementarity between traditional microbiological methods and microbial genomics for monitoring circulating 7PET strains and tracking their evolution development and that of AMR in particular.

**IMPACT STATEMENT:** The number of cholera cases has fallen since the beginning of 2024 (only 49 cases reported up to July 28), but Cameroon, in Central Africa, experienced a number of cholera epidemics between 2018 and 2023, with 26,411 cholera cases in total and 735 deaths (case fatality rate: 2.8%). We performed a phenotypic and genomic analysis of 164 *V. cholerae* O1 isolates representative of the successive outbreaks that were collected from 2018 to 2023 from seven of the 10 regions of Cameroon, which revealed that these isolates were very similar and closely related to other West African isolates. All belonged to the same pandemic lineage thought to have been introduced into West Africa from South Asia in 2007. The strains of this lineage have become less resistant to antimicrobial drugs over time, due to a single genetic event (i.e., deletion of a region of the bacterial chromosome containing several antimicrobial resistance genes) and have reverted to the Ogawa serotype (the original serotype of wild-type strains). Neither of these changes were anticipated. Furthermore, our findings support the hypothesis that the causal agent of cholera is permanently circulating in the countries around the Lake Chad basin and those of West Africa. They stress the importance of combining a genomic approach with traditional microbiology for the epidemiological surveillance of cholera. Furthermore, transborder collaborative efforts are strongly advised to ensure effective cholera control in this region.

## INTRODUCTION

Cholera is an acute diarrhoeal infection of humans caused by toxigenic *Vibrio cholerae* O1 or O139 bacteria. It spreads via the faecal-oral route and occurs following the ingestion of contaminated water or food or through direct contact with the body fluids or body parts of infected humans (1). Cholera is primarily linked to poor access to safe water and inadequate sanitation, and it can spread rapidly, causing epidemics and pandemics. The pathogenicity of this bacterium results from its ability to produce cholera toxin (CT) and toxin co-regulated pili (TCP) in the human intestine. Cholera can lead to death within hours if left untreated (2). Its treatment involves the oral administration of saline rehydration solutions or the intravenous administration of fluids, depending on disease severity. Antibiotics can also be administered as a supplementary treatment in severely dehydrated patients (3). The choice of antibiotic depends on the susceptibility profile of the circulating *V. cholerae* strain. The Global Task Force on Cholera Control (GTFCC) recommends tetracyclines, fluoroquinolones and macrolides as the antibiotics of choice for cholera treatment. Doxycycline should be the first choice, but ciprofloxacin and azithromycin can be used as alternatives. Antibiotic therapy helps to reduce the duration of diarrhoea, *V. cholerae* excretion and, consequently, cholera transmission (4).

Since the early 1960s, the world has been in the grip of the seventh pandemic of cholera caused by *V. cholerae* serogroup O1 of the El Tor biotype (5). Cholera has become endemic in a number of geographic areas, reflecting a failure to implement adequate control measures. Its spread was intensified by conflicts, mass displacements, disasters from natural hazards and climate change (6). This disease remains a major public health concern, especially in Asia and in sub-Saharan Africa (7). In 2022, the World Health Organization (WHO) reported an upsurge in the number of cholera cases, with an increase in the number of countries reporting cases, including reports of very large outbreaks (>□10,000 suspected cases) in seven countries on two continents (Afghanistan, Cameroon, Democratic Republic of the Congo, Malawi, Nigeria, Somalia and Syria) (8). In January 2023, the WHO even classified this global resurgence of cholera as a grade 3 emergency, the highest level of emergency accorded (9).

Cameroon is a lower middle-income country with a population of over 27.9 million (2022) people. This state, covering 475,442 km², is located in Central Africa with an Atlantic coast and shared five borders with the Central African Republic, Chad, Equatorial Guinea, Gabon, and Nigeria (10). Cameroon, like several other countries in West and Central Africa, has been experiencing recurrent cholera outbreaks (11,12). Six months after cholera hit West Africa (in Guinea) in August 1970, the first cases appeared in Douala – the largest city in Cameroon and home to Central Africa’s largest port – in February 1971. Between 1971 and 1990, the number of cholera cases reported to the WHO appeared relatively low (a total of 5,130 cases, including 2,157 cases in 1971 and 1,158 cases in 1985). This may reflect underreporting — for example 55 cases were reported in 1983, whereas Garrigue *et al.* (13) mentioned 4,423 cholera cases with 119 deaths in Douala this same year — or even no reporting at all for some years (Table S1). From 1991, the number of reported cases increased sharply, with a total of 96,234 cases between 1991 and 2023. The number of cases annually has increased most steeply since 2010, with 10,759 cases (and 657 deaths) that year, 22,433 cases (and 783 deaths) in 2011, and 14,431 cases (and 279 deaths) in 2022. By contrast, no cholera cases were identified at all in several years (for example, in the recent past, in 2008, 2016 and 2017). After a lull for most of 2021, probably due to the implementation of preventive measures for COVID-19, such as the promotion of hand-washing and hygiene, social distancing and the banning of large gatherings (14), a large cholera outbreak began in October 2021, after the identification of two cases in the Ekondo Titi health district (Southwest region). Epidemiological data showed that the majority of cholera cases were notified in 16 active health districts in four active regions (Littoral, Centre, South and West) in April 2023, particularly during the rainy season (15).

Genomic studies showed that the seventh pandemic *V. cholerae* O1 strains belonged to a particular lineage, 7PET, which has spread by periodic radiation from a single source population located in the Bay of Bengal (South Asia) (16). The 7PET lineage was introduced into Africa at least 11 times between 1970 — when the seventh pandemic first hit Africa — and 2014 (sublineages AFR1, AFR3-AFR12, formerly T1, T3-T12) (17,18). These introductions were followed by clear regional transmission crossing into bordering countries (19). At least three new introductions were subsequently identified in Eastern Africa (7PET sublineages AFR13 and AFR15) (20,21) and Algeria (sublineage AFR14) (22), all following the same Asia-to-Africa introduction pattern.

In Cameroon, four 7PET sublineages (AFR1, AFR7, AFR9, and AFR12) were identified during the cholera outbreaks occurring between 1971 and 2019 (17,19,23). The most recently introduced sublineage, AFR12 (first isolates detected in 2009) was identified during a severe cholera outbreak in 2010-2011 (∼34,000 cases and ∼1,500 deaths) and in neighbouring countries in West and Central Africa, such as Nigeria in 2010 (44,456 cases and 1,712 deaths), 2014 (35,996 cases and 755 deaths) and 2018 (45,037 cases and 836 deaths) (17,19).

We used conventional microbiology and microbial genomics to characterise the virulence and antimicrobial resistance (AMR) determinants of 164 clinical isolates of *V. cholerae* O1 collected during cholera outbreaks in Cameroon from 2018 to 2023 and representative of the successive outbreaks. Our phylogenomic analysis on these and other genomes (> 1,700 7PET genomes in total) also aimed to determine whether the strains responsible for the recent increase in cholera cases in Cameroon since October 2021 still belong to sublineage AFR12 or to a newly introduced 7PET sublineage.

## METHODS

### Microbiological analysis and data collection

Microbiological analysis was performed on stool samples from patients with suspected cholera at the bacteriology laboratory of the *Centre Pasteur du Cameroun* (CPC) in Yaoundé (one of the reference laboratories for cholera in Cameroon) and the CPC antenna in Garoua, as previously described (24). Briefly, in cases of positive culture on thiosulphate-citrate-bile-saccharose agar (TCBS) (Deben Diagnostics, Ipswich, United Kingdom), purified colonies were identified with biochemical tests and then serogrouped and serotyped by the sero-agglutination method (Denka Seiken, Tokyo, Japan). In total, 164 *V. cholerae* O1 isolates collected during cholera outbreaks in Cameroon between 2018 and 2023 were recovered and studied. Of these 164 isolates, 35 were collected in 2018, 28 in 2019, 41 in 2020, 4 in 2021, 39 in 2022, and 17 in 2023 (Table S2). These isolates were also sent to the French National Reference Centre for Vibrios and Cholera (FNRCVC), *Institut Pasteur* (IP), Paris, France, for further characterisation.

### Antimicrobial susceptibility testing

Antimicrobial drug susceptibility testing was performed by the disk diffusion (DD) method on Mueller-Hinton agar (Bio-Rad, Marnes-la-Coquette, France) according to the guidelines of the European Committee on Antimicrobial Susceptibility Testing (EUCAST) (https://www.eucast.org/ast_of_bacteria). The following disks (Bio-Rad) were used for the DD method: ampicillin (AMP, 10 µg), cefotaxime (CTX, 30 µg then 5 µg), meropenem (MEM, 10 µg), azithromycin (AZM, 15 µg), sulphonamides (SUL, 200 µg), O/129 vibriostatic agent, (O129, 500 µg) or trimethoprim (TMP, 5µg), trimethoprim-sulfamethoxazole (SXT, 1.25 µg/23.75 µg), chloramphenicol (CHL, 30 µg), nitrofurantoin (NIT, 300 µg, then 100 µg), tetracycline (TET, 30 µg), doxycycline (DOX, 30 µg), minocycline (MIN, 30 µg), and polymyxin B (PXB, 300 UI). The isolates were tested for their susceptibility to nalidixic acid (NAL) and ciprofloxacin (CIP) by either the microdilution method (Sensititre^TM^ Thermo Fisher Scientific, Cleveland, OH, USA) or E-tests (bioMérieux, Marcy L’Etoile, France). The EUCAST criteria for the interpretation of antimicrobial drug susceptibility testing results for *Vibrio* spp. (v. 14.0) were used when available (25). In particular, resistance to CIP was defined as a minimum inhibition concentration (MIC) > 0.25 mg/L. However, as a means of distinguishing isolates susceptible to CIP (MIC□≤□0.25□mg/L) that are wild-type (WT) from those that are non-WT, we defined two categories based on epidemiological cutoffs (ECOFFs): decreased susceptibility to ciprofloxacin (MIC between 0.03 and 0.25□mg/L) and true susceptibility to ciprofloxacin (MIC□≤□0.016□mg/L). For AMP, CTX (when 30 µg disks were used), SUL, and CHL, the Clinical and Laboratory Standards Institute (CLSI) interpretative criteria for *Vibrio* spp. were used (26). For NIT, MIN, and NAL, the CLSI interpretative criteria for Enterobacterales were used (27). For O129 and PXB, we defined susceptibility as an inhibition zone diameter ≥ 15 mm (and resistance as an inhibition zone diameter < 15 mm). *Escherichia coli* ATCC^®^ 25922™ was used for internal quality control.

### Total DNA extraction and whole-genome sequencing

Genomic DNA was extracted from pure cultures of *V. cholerae* O1 on alkaline nutrient agar (20 g casein meat peptone E2 [Organotechnie, La Courneuve, France]; 5 g sodium chloride [Sigma, St. Louis, MO, USA]; 15 g Bacto agar [BD Difco, Franklin Lakes, NJ, USA]; distilled water to 1LL; adjusted to pH 8.4; autoclaved at 121 °C for 15 min), at FNRCVC, with the Promega Maxwell 16 Cell DNA Purification Kit on a Maxwell 16 instrument (Promega, Madison, WI, USA), in accordance with the manufacturer’s instructions. Whole-genome sequencing was performed at the genotyping and sequencing core facility of the *Institut du Cerveau*, Paris, France (https://institutducerveau.org/) on Illumina platforms (NovaSeq6000 or NovaSeq X Plus) generating 151 bp paired-end reads; yielding a mean coverage of 548-fold (minimum 167-fold, maximum 895-fold).

### Genomic sequence analyses

After Illumina sequencing, short reads were filtered with FqCleanER v.23.07 (https://gitlab.pasteur.fr/GIPhy/fqCleanER) to eliminate adaptor sequences and discard low-quality reads with phred scores□<□28 and a length□<□70□bp. Read quality was assessed with FastQC v.0.11.9 (https://github.com/s-andrews/FastQC). Taxonomic read classification with Kraken v.2.1.13 was used to confirm that sequencing reads originated from *V. cholerae* and not from a contaminant (28). Only genomes satisfying the quality control criteria were retained for further analyses. SPAdes v.3.15.5 (29) was used to assemble genomes. Sequence type (ST) determination was performed with the multilocus sequence type (MLST) scheme of Octavia *et al.* (17). Different genetic markers were analysed with BLAST v.2.2.26 against reference sequences for the O1-antigen biosynthesis gene *rfb*, *tcpA,* the major pilin subunit of the toxin-coregulated pilus (TCP), *ctxB* encoding the B subunit of cholera toxin (CT)*, wbeT,* which has been implicated in the Ogawa/Inaba serotype shift, and *Vibrio* pathogenicity islands (VPI-1, VPI-2, VSP-I, and VSP-II), as previously described (17). The presence and type of acquired antimicrobial resistance genes (ARGs) or ARG-containing structures were determined with ResFinder v.4.0.1 (https://cge.food.dtu.dk/services/ResFinder/) (30), BLAST analysis against SXT/R391 integrative and conjugative elements, and PlasmidFinder v.2.1.1 (https://cge.food.dtu.dk/services/PlasmidFinder/) (31). We checked for mutations of genes encoding resistance to quinolones (*gyrA*, *parC*), resistance to nitrofurans (*nfsA*, nitroreductase, VC0715 and VCA0637, dihydropteridine reductase), or restoring susceptibility to polymyxin B (VC1320, *vprA*), by performing a manual analysis of the sequences assembled *de novo* with BLAST, as previously described (20).

### Phylogenetic analysis

Repetitive (insertion sequences and the TLC-RS1-CTX region) and recombinogenic (VSP-II) regions in the alignment were masked (17). Putative recombinogenic regions were detected and masked with Gubbins v.3.2.0 (32). A maximum-likelihood (ML) phylogenetic tree was built from an alignment of 10,339 chromosomal SNVs, with RAxML version 8.2.12, under the GTR model with 200 bootstraps (33). This global tree was rooted on the A6 genome (the earliest and most ancestral seventh pandemic isolates) collected in Indonesia in 1957 (17) and visualised with Interactive Tree of Life (iTOL) version 6 (https://itol.embl.de) (34).

## RESULTS

### Spatiotemporal distribution and characteristics of the *V. cholerae* O1 isolates studied

We studied 164 *V. cholerae* O1 isolates collected in Cameroon between 2018 and 2023 (Fig. 1A and 1B, Table S2). The sampling area was known for 161 of these isolates, which originated from seven of the 10 regions of Cameroon: North (65/160; 40.6%), Centre (56/160; 35%), Littoral (16/160; 10%), South (21/160; 13.12%), Adamawa (1/160; 0.62%), Southwest (1/160; 0.62%), and Northwest (1/160; 0.62%) (Fig. 1A, Table S2). The two serotypes of *V. cholerae* O1 (i.e., Inaba and Ogawa) were observed during the study period, with 61.6% (*n* = 101) of isolates belonging to Inaba and 38.4% (*n* = 63) to Ogawa (Fig. 1C). A shift between these two serotypes occurred in 2021. All but three of the 104 isolates collected between 2018 and 2020 belonged to Inaba, whereas Ogawa was the only serotype identified for the 60 isolates collected between 2021 and 2023 (Fig. 1C).

**Figure 1.**
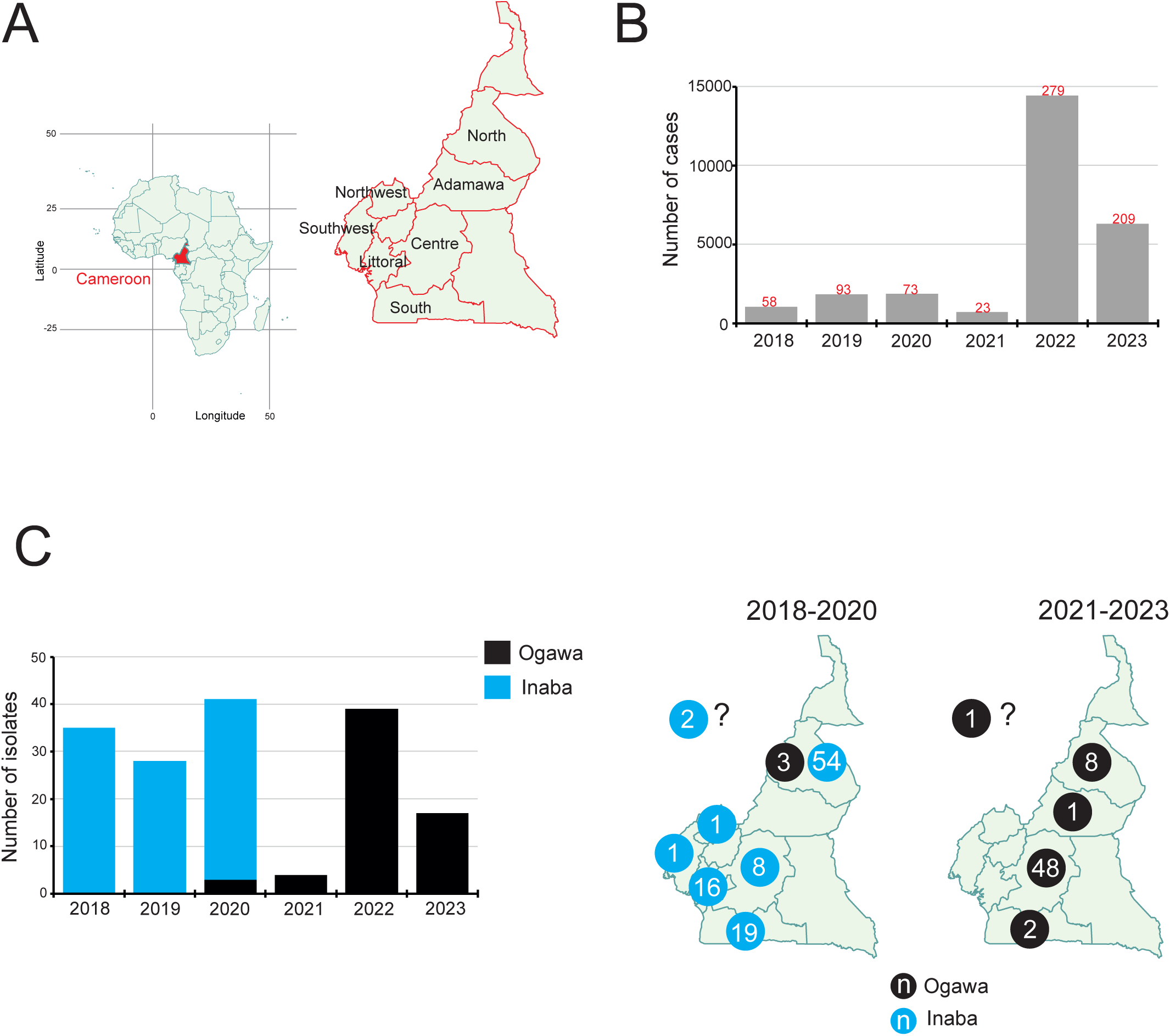
Cholera cases and *Vibrio cholerae* O1 isolates studied, Cameroon, 2018-2023. **(A)** On the left, location of Cameroon on the African continent. This map was drawn with the R package “maps”. On the right, a magnification of Cameroon showing the 10 administrative regions. This map is a modified version of the cm#admin1 image (https://simplemaps.com/svg/country/cm#admin1), created by simplemaps (https://simplemaps.com/) and freely available for commercial and personal use. The seven regions from which *V. cholerae* O1 isolates were collected for our study are indicated. Douala, the economic capital of Cameroon, is the capital of the Littoral region and Yaoundé, the capital city of Cameroon, is also the capital of the Centre region. (**B)** Number of cholera cases reported by Cameroon to the World Health Organisation between 2018 and 2023. The numbers of reported deaths from cholera are indicated in red above the bars. **(C)** On the left, temporal distribution of the 164 *V. cholerae* O1 isolates studied here, by serotype (Ogawa vs. Inaba). On the right, geographic distribution (at regional level) of the 164 *V. cholerae* O1 isolates, by serotype (Ogawa vs. Inaba). “*n*” indicates the number of isolates per serotype. The question mark indicates that the region of isolation is unknown.

**Figure 2.**
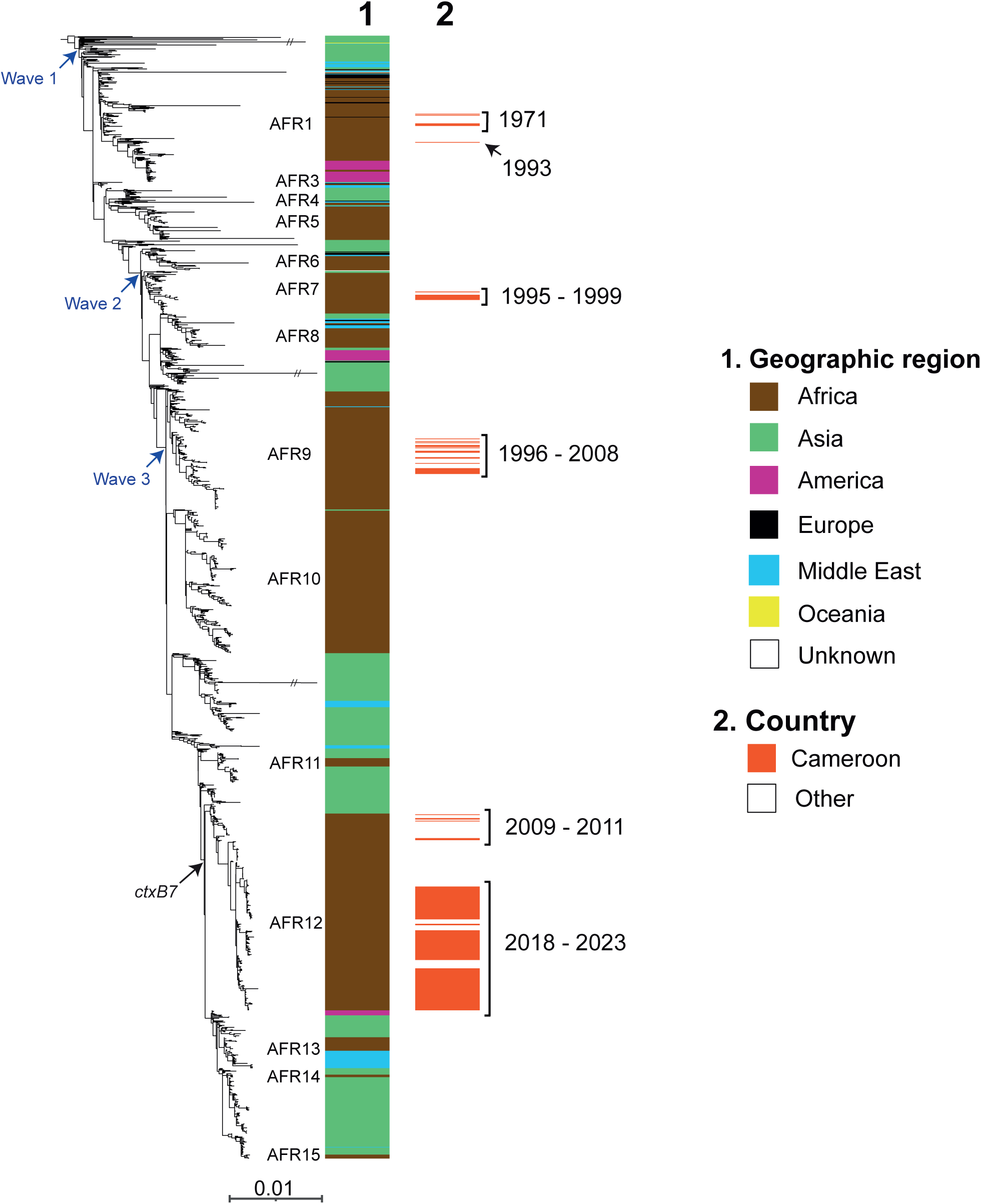
Maximum-likelihood phylogeny of 1,742 seventh pandemic *Vibrio cholerae* O1 El Tor isolates, including 164 collected in Cameroon in 2018-2023. A6 was used as the outgroup. Blue arrows represent the three genomic waves and the black arrow indicates the acquisition of the *ctxB7* allele, the most recent variant of the gene encoding the B subunit of cholera toxin. The colour coding in the first column shows the geographic origins of the isolates, and African sublineages (AFR1, AFR3–AFR15) are shown on the left. The red colour in the second column indicates the Cameroonian origin of the isolates (164 isolates studied collected in 2018-2023 and 45 previously published and collected between 1971 and 2011). The years of isolation of Cameroonian *V. cholerae* O1 isolates are indicated at the right of the second column. Double slashes indicate an artificial shortening of the branch to enhance visualisation. Scale bars indicate the number of nucleotide substitutions per variable site.

### Genomic features of the *V. cholerae* O1 isolates studied

All 164 *V. cholerae* O1 isolates belonged to the 7PET lineage according to their multilocus sequence type (ST69). They all carried the *ctxB7* allele (characterised by a 1-nt variant of codon 19 resulting in replacement of the classical cholera toxin B histidine residue with asparagine) as found in the most recent genomic wave 3 7PET isolates. They also harboured the toxin-coregulated pilus subunit A gene variant *tcpA*^CIRS101^, and a deletion encompassing VC_0495 to VC_0512 (according to GenBank accession no. AE003852) in the *Vibrio* seventh pandemic island II (VSP-II) (Table S3).

All 101 *V. cholerae* O1 serotype Inaba isolates had a point mutation (G674A) in the *wbeT* (formerly *rfbT*) gene, which encodes a methyltransferase. This alteration, named alteration C19 by Weill *et al*. (17), encodes the C225Y amino-acid substitution leading to the shift in serotype from Ogawa to Inaba. The 63 *V. cholerae* O1 serotype Ogawa isolates did not carry this alteration to *wbeT*.

The 164 *V. cholerae* O1 isolates contained the *dfrA1* gene encoding resistance to trimethoprim and to the O/129 vibriostatic agent (Table S4). This gene was located on ICE*Vch*Ind5, a member of the SXT-R391 SXT family of integrative and conjugative elements (ICE). However, this genomic element included a deletion of about 10 kb (encompassing the sequence from ICEVCHIND5_0011 to ICEVCHIND5_0021 according to GenBank accession no. GQ463142) in variable region III, resulting in the loss of the other four AMR genes encoding resistance to streptomycin (*strA* and *strB*), sulphonamides (*sul2*), and chloramphenicol (*floR*) (22,23). All 164 *V. cholerae* O1 isolates carried chromosomal point mutations (i) in VC_0715 (*nfsA*), resulting in the R169C substitution, and in VC_A0637 (*nfsB*, resulting in the premature stop codon (Q5Stop), conferring nitrofurantoin resistance; (ii) in the DNA gyrase gene, *gyrA* (S83I), and in the topoisomerase IV gene, *parC* (S85L), conferring resistance to quinolones. By contrast, none of the 164 isolates had the C505T mutation in the *vprA* gene (VC_1320) associated with a reversal of susceptibility to polymyxins seen in sublineages AFR13 to AFR15. This genomic AMR profile was concordant with the phenotypic resistance profile obtained from a subsample of 99 isolates resistant to the O/129 vibriostatic agent (equivalent to trimethoprim) or to trimethoprim, nitrofurantoin, nalidixic acid with either decreased susceptibility to ciprofloxacin (MIC of 0.25 mg/L) or resistance to ciprofloxacin (MIC > 0.25 mg/L), and to polymyxin B (Table S2). As predicted by genomic analysis, these isolates remained susceptible to other antimicrobial drugs recommended for cholera treatment, such as tetracyclines (first-line treatment) and azithromycin (alternative treatment) (35).

### Phylogenomic analysis of the *V. cholerae* O1 isolates studied

The 164 *V. cholerae* O1 isolates collected between 2018 and 2023 in Cameroon clustered together within the AFR12 sublineage (Fig. S1). They were closely related to isolates from this sublineage collected in Niger, Benin and Togo — neighbouring countries — between 2018 and 2020 (Fig. S1). The Cameroonian isolates collected in 2009-2011 also belonged to sublineage AFR12, but were located on more ancestral branches of this sublineage. Other published older Cameroonian isolates (collected from 1971 to 2008) were found to belong to the various sublineages (AFR1, AFR7, and AFR9) identified in West Africa during the seventh cholera pandemic. Interestingly, the recent Ogawa isolates (identified since 2020) were not of clonal origin but derived from two different Inaba clades. The larger group of isolates (*n* = 51) with a reversion to the Ogawa serotype came from the Centre region of Cameroon and was collected between 2021 and 2023. These isolates were closely related to serotype Ogawa isolates collected in Togo in 2020, which were, in turn, closely related to serotype Inaba isolates collected in Benin in 2019. The smaller group (*n* = 12) with a reversion to serotype Ogawa was collected in the North region of Cameroon between 2020 and 2022 and these isolates were closely related to serotype Inaba isolates collected in the same region (North) in 2019.

## DISCUSSION

Based on the available genomic sequences for *V. cholerae* O1 strains isolated in Cameroon since 1971, the year in which the seventh pandemic of cholera hit this country, four different 7PET sublineages (AFR1, AFR7, AFR9, and AFR12) were identified from the three genomic waves described. Other than during the 1996-1999 period (when two sublineages, AFR7 and AFR9, were identified), only one sublineage was found at a time. The AFR9 sublineage, which belongs to the earliest genomic wave 3 group (carrying the *ctxB1* allele) was last detected in Cameroon in 2008 and in West Africa in 2012 (in Guinea and Guinea Bissau). However, these observations are based on the limited number of Cameroonian bacterial genomes available. Our genomic study was performed on the largest dataset to date (*n* = 164) for *V. cholerae* O1 isolates from seven different regions collected over a six-year period. Even though our sampling encompasses only 0.6% of the cholera cases reported during the study period, the sample size of this study and its spatiotemporal representativeness support the hypothesis that the AFR12 sublineage — which probably emerged between 2007 and 2009 — was the only sublineage circulating in Cameroon during the 2018-2023 period. In 2022, the European countries reported 51 cholera cases, including five linked to Cameroon (two cases infected in Cameroon and three cases infected in Europe after consumption of food imported from Cameroon). All five isolates belonged to the AFR12 sublineage (36). A recent study published during the drafting of this manuscript described 13 genomic sequences from *V. cholerae* O1 isolates isolated in Cameroon between 2020 and 2023, all from the AFR12 sublineage (37). Similarly, a preprint (38) released during the drafting of this manuscript reported one *V. cholerae* O1 genomic sequence from Cameroon in 2020 and 30 in 2023, all belonging to AFR12. The AFR12 sublineage belongs to the most recent wave 3 group first identified in Southern Asia in the early 2000s before spreading globally, reaching Haiti in 2010. Such wave 3 strains with the *ctxB7* allele have been shown to have a phenotypic profile compatible with hypervirulence (e.g. enhanced toxin and haemolysin production, hypermotility and greater fitness during colonisation and lethality in infant mice) (39).

The significant increase in the number of reported cholera cases in Cameroon in 2022-2023 (total of 20,901 cases) is therefore not due to the introduction of a new sublineage. This increase occurred at the same time as a reversion from serotype Inaba to serotype Ogawa. Hence, from 2021 to the end of the study period in 2023, all the isolates studied were of serotype Ogawa. This emergence of serotype Ogawa isolates was not due to a clonal expansion of a single bacterial population. Instead, it was due to two different clades, the first found in Central Cameroon from 2021 to 2023 and the second in North Cameroon from 2020 onwards. This reversion from Inaba to Ogawa is not a new phenomenon (40,41), as three different reversion events have already been reported in the AFR9 sublineage (17). One of these events occurred in Cameroonian isolates. In Cameroon, from 1996 to 2005, all AFR9 7PET isolates were of serotype Inaba, whereas, from 2006 to 2008, all isolates were of serotype Ogawa. The *wbeT* alteration (G674A) leading to the Inaba phenotype was also identical in both the AFR9 and AFR12 sublineages. This rare alteration had previously been observed only in the AFR9 sublineage, despite the analysis of 1,070 global 7PET genomes (17).

The switch from the Ogawa to the Inaba serotype seems to occur readily in *V. cholerae* O1, both *in vivo* and *in vitro* (41,42). A reciprocal interconversion between these two serotypes has been reported in germ-free mice treated with anti-Ogawa serum (43). It has been suggested that this phenomenon occurs naturally during cholera infection, due to either a spontaneous mutation or the host immune response (43,44). The presence of agglutinating antibodies specific to a particular serotype in the serum may exert a powerful selective effect on the growth of other *Vibrio* serotypes (41,42,45). This interpretation is consistent with the results of an *in vitro* study by Sakazaki *et al*. in 1971, which reported the selection of the Inaba variant with anti-Ogawa sera (46). As already suggested for regions in which cholera is endemic (47), we can hypothesise that switches in serotype have contributed to the persistence of cholera in Cameroon.

A recent study by Ngomtcho *et al.* (37) in which antimicrobial drug susceptibility was assessed by the disk diffusion method reported surprisingly high rates of resistance to azithromycin (49% of isolates), doxycycline (54%), and third-generation cephalosporins (28%) in 54 *V. cholerae* O1 isolates from Cameroon collected between 2019 and 2023. However, an *in silico* analysis of 13 sequenced isolates identified no AMR genes that could explain this very unusual resistance phenotype that had never previously been reported in the region. Mboowa *et al.* (37), reported the prediction of AMR genes from genomic sequences but did not perform any antimicrobial drug susceptibility testing to confirm their predictions. We performed standardized antimicrobial drug susceptibility testing on almost one hundred isolates collected from the same country over the same period, and we did not observe this trend in antimicrobial drug resistance. All the isolates tested were resistant only to nitrofurans, trimethoprim, and colistin and had decreased susceptibility to ciprofloxacin (most — 65.7%, 65/99 — being classified as resistant to ciprofloxacin based on clinical breakpoints). These phenotypic antimicrobial drug susceptibility data were consistent with genomic predictions of AMR. Interestingly, the 164 AFR12 *V. cholerae* O1 isolates collected between 2018 and 2023 in Cameroon were less resistant to antimicrobial drugs than the AFR12 isolates collected in the same country during the 2009-2011 period. All AFR12 isolates since 2009 have been found to carry *gyrA* and *parC* mutations decreasing susceptibility or even conferring resistance to ciprofloxacin, whereas all 164 recent isolates had a deleted form of the ICE*Vch*Ind5 genomic element resulting in the loss of four AMR genes (*strA*, *strB*, *floR* and *sul2*). These AMR genes are no longer under selection pressure as they encode resistance to drugs not currently recommended for the treatment of cholera. This deletion, mediated by two transposable elements, occurred sporadically in the upper part of the AFR12 phylogenetic tree, subsequently concerning all isolates, with a shift from the Ogawa to the Inaba serotype from 2018.

AFR9 *V. cholerae* O1 isolates previously displayed multidrug resistance (MDR) due to the presence of ICE*Vch*Ind5 but were susceptible to quinolones (wild-type *gyrA* and *parC* genes). Older AFR7 and AFR1 genomes had no AMR genes, with the exception of F135 (AFR1 sublineage), which was isolated in Cameroon in 1993 and contained an IncC plasmid conferring MDR. F135 contained *bla*_CARB-4_ (encoding a carbenicillin*-*hydrolysing beta-lactamase), *tet(C)* (conferring resistance to tetracycline), *strA* and *strB* (resistance to streptomycin), *aac(6’)-IIc* (resistance to gentamicin), *sul1* and *sul2* (resistance to sulphonamides), *dfrA15* (resistance to trimethoprim and the vibriostatic compound O/129, and to cotrimoxazole in combination with a *sul* gene), and *catA1* (resistance to chloramphenicol). This genome is probably a relic of the MDR *V. cholerae* O1 isolates identified in Douala in 1984-1985 following massive chemoprophylaxis with sulphadoxine in April 1983 (in response to a large cholera outbreak, with 4,423 cholera cases, that occurred in Douala in 1983) (13). These 1984-1985 isolates were reported to have AMR profiles (13) consistent with that of F135 and a molecular study on some of these 1984-1985 isolates performed in 1998 identified a non-TEM-1/non-OXA-1 beta-lactamase and a new type of *dfr* gene (48), these findings also being consistent with the AMR gene content of F135 (*bla*_CARB-4_ and *dfrA15* genes).

Since 2018, highly-drug resistant 7PET isolates containing MDR IncC plasmids have increasingly been detected in the Middle East and Eastern Africa (49–51). These isolates of concern (as they are resistant to ciprofloxacin, in addition to tetracyclines or azithromycin) belong to the AFR13 sublineage, which is also part of the most recent genomic wave 3 group (i.e. with the *ctxB7* allele). These AFR13 isolates also had the ICE*Vch*Ind5 form with a 10 kb deletion and carried the same *gyrA* and *parC* mutations as AFR12 isolates. Fortunately, no MDR IncC plasmids were identified in our 164 *V. cholerae* O1 isolates.

This study provides support for the idea that, in tandem with traditional microbiological methods, microbial genomics is a powerful tool for monitoring circulating 7PET strains and tracking their development, particularly as concerns the development of AMR. Nevertheless, standardised antimicrobial susceptibility testing remains useful for the regular monitoring of *V. cholerae* O1 outbreak isolates and the identification of new molecular mechanisms of antimicrobial resistance, which may arise periodically, albeit rarely. The identification of unusual phenotypes of resistance to particular antibiotic classes by one laboratory, particularly if these findings are not supported by the corresponding AMR gene content of the genome, should lead to careful assessment of the antimicrobial drug susceptibility testing process. With the increasing use of purely genomic studies, a validation of the different AMR prediction tools will be also important, as genes falsely predicted to be associated with AMR, such as *catB9* and chloramphenicol resistance (17) (and probably also *varG* and carbapenems and *almEFG* and polymyxins), are frequently reported (37, 38).

Finally, microbial genomics is an unequaled tool for studying the regional circulation of the 7PET agent via real-time transborder monitoring. The dynamics of cholera epidemics should be investigated not only at national level but also at regional level due to the influence of cross-border epidemics, particularly around the Lake Chad basin. Between 2010 and 2012, the epidemiological landscape in neighbouring countries (Nigeria, Chad and Niger) was similar to that in Cameroon; the epidemic peaked in Nigeria (44,456 cases) in 2010, in Chad (17,267 cases) and Cameroon (22,433 cases) in 2011, and then in neighboring Niger (5,284 cases) in 2012 (17). This study was not designed to use a better granulometry to understand the regional spread of AFR12, even though we included contemporary bacterial genomes from other West African countries, such as Benin, Togo, and Niger. However, we are confident that the regular release of new 7PET genomic sequences from the region, particularly from Nigeria, a country heavily affected by the disease (e.g. the 131 genomes recently released in the preprint by Mboowa *et al*. (38)), will, in the near future, provide a comprehensive regional genomic framework for the 7PET agent, supporting research studies aiming to improve our understanding of the principal factors driving cholera outbreaks in West and Central Africa.

## Data Availability

All data produced in the present work are contained in the manuscript

## Data availability

The short-read sequence data generated in this study have been submitted to the European Nucleotide Archive (ENA, http://www.ebi.ac.uk/ena). Their individual accession numbers are listed in Table S5.

All the accession numbers of the publicly available sequences – downloaded from GenBank (https://www.ncbi.nlm.nih.gov/genbank/) or ENA – and used in this study are listed in Table S5.

## Acknowledgements

We would like to thank all the staff and personnel of the laboratories participating in this study including the team of the bacteriology laboratory of the *Centre Pasteur du Cameroun* in Yaoundé and the Garoua annexe; the team of the *Unité des Bactéries pathogènes entériques*, and the International Direction of the *Institut Pasteur* in Paris. Special thanks also to Sara Eyangoh and Marianne Lucas-Hourani, Lise Frézal, Magali Lago, Sylvie Guillemaut, Maud Seguy, Thierry Planchenault, Paul Martin and Quentin Holleville.

## AUTHOR CONTRIBUTIONS

Conceptualisation: F.-X.W., J.-M.C., P.L.S.T; Methodology: P.L.S.T, F.X.W.; Investigation and resources: P.L.S.T., R.M.N, E.N., L.D.T.M., W.M, M.A, E.S., L.N.-J., F.T., M.A., C.T.R.F., U.N., A.O., R.P., C.R., M.-L.Q. Y.E.D.D., H.B.S., H.A.W., A.Y.S., F.S.; Validation: P.L.S.T., F.X.W.; Data curation: P.L.S.T., E.N., M.-L.Q., F.X.W; Writing – original draft preparation: P.L.S.T, E.N., J.-M.C., F.-X.W.; Writing – critical review: A.N., M.-L.Q., R. P., C.R. All the authors have read and agreed to publication of the submitted version of the manuscript.

## ETHIC OVERSIGHT

This study was based exclusively on bacterial isolates and associated metadata collected under the mandate for laboratory-based surveillance of cholera awarded by the Ministry of Health of Cameroon to the Centre Pasteur du Cameroun. The associated metadata did not contain any personal identifiable information and were restricted to year and province of isolation of the *V. cholerae* O1 isolates. As a result, neither informed consent nor approval from an ethics committee was sought given this is not research conducted on human participants.

---

**Fig. S1.**
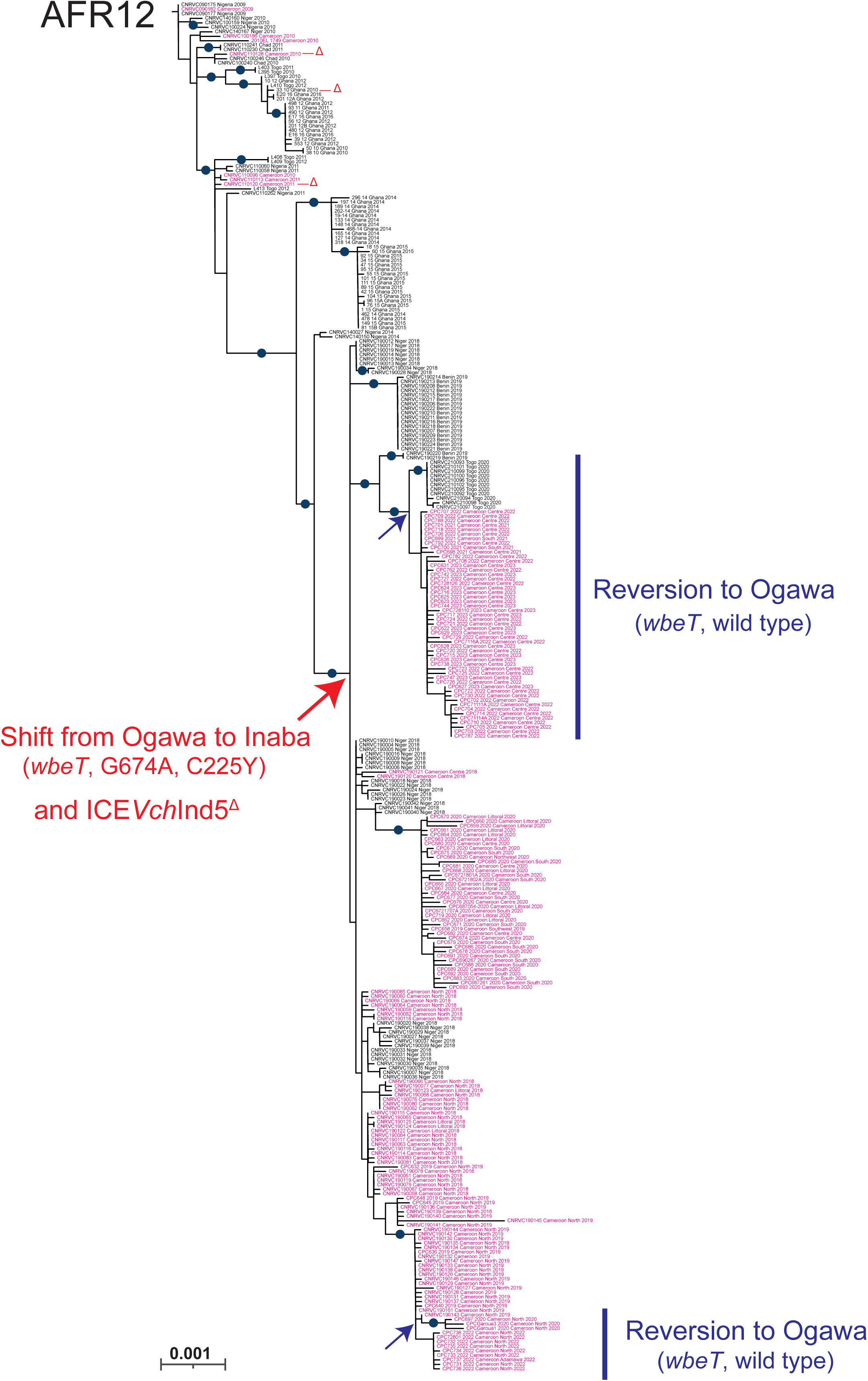
Maximum-likelihood phylogeny of 305 *Vibrio cholerae* O1 El Tor isolates from the AFR12 sublineage, including 164 collected in Cameroon in 2018-2023. This figure is a magnification of the branch for the AFR12 sublineage presented in Fig. 2. For each genome, its name (or accession number), the country in which contamination occurred (and the region for Cameroonian isolates) and the year of sample collection are indicated at the tip of the branch. The Cameroonian isolates are shown in pink. The red arrow indicates the acquisition of the *wbeT* mutation G674A (encoding amino-acid substitution C225Y) associated with the Ogawa-to-Inaba serotype shift and a deleted form of the ICE*Vch*Ind5 genomic element encoding antimicrobial resistance. Purple arrows indicate a reversion to the Ogawa serotype. The sporadic acquisition of deleted variants of ICE*Vch*Ind5 is indicated by a red Δ. Scale bars indicate the number of nucleotide substitutions per variable site. Blue circles correspond to bootstrap values□≥□95%.

